# Distinct and synergistic immunological responses to SARS-CoV-2 and *Mycobacterium tuberculosis* during co-infection identified by single-cell-RNA-seq

**DOI:** 10.1101/2023.05.24.23290499

**Authors:** Dylan Sheerin, Thanh Kha Phan, Emily M. Eriksson, COVID PROFILE Consortium, Anna K. Coussens

## Abstract

COVID-19 and tuberculosis (TB) exhibit similar symptomatic presentation, clinical parameters and co-diagnosis increases COVID-19 mortality yet there is limited understanding of the mechanisms underlying their immunopathogenic interactions. Here we show by single-cell RNA-sequencing of 18,990 cells from whole blood uninfected or infected with *Mycobacterium tuberculosis* (*Mtb*), SARS-CoV-2, or both pathogens, their shared, distinct, and synergistic immunological interactions. The greatest transcriptional divergence occurred within monocytes and two neutrophil subsets at early timepoints of infection. Co-infection had the greatest synergistic effect 24 hours post-infection including enrichment of IFN-γ and TNF production, whilst 96 hours post-infection *Mtb*, SARS-CoV-2 and co-infection shared considerable pathway overlap. SARS-CoV-2 infection alone resulted in widespread cell death 96 hours post-infection, whilst *Mtb* and co-infection had enhanced cell survival at 96 hrs, sharing negative regulation of extrinsic apoptosis. Our findings elucidate potential pathways for targeted host-directed therapies, which is particularly crucial for settings where these pathogens are now endemic.

## Introduction

The COVID-19 pandemic has had a substantial impact on global progress towards tuberculosis (TB) elimination, primarily through disruptions to TB diagnosis and treatment services^1, 2^. There was a precipitous drop in TB notifications in 2020 and, according to numbers derived from the most recent World Health Organization report, annual deaths from TB have increased between 2019 and 2021 for the first time since 2005^3^, reversing a trend of slow but sustained decline. Beyond impacting TB health services, the Global Tuberculosis Network conducted a prospective multi-country register-based cohort study involving 175 centres across 37 countries, identifying 767 dually diagnosed TB/COVID-19 patients which provided evidence that TB patients suffer worse outcomes when co-infected by COVID-19^4^. They found that TB/COVID-19 patients experienced 11.1% mortality, compared to 1-2% for COVID-19 alone. A large multi-variate analysis of 3,460,932 patients in South Africa found that current and previous TB increased COVID-19 mortality with adjusted hazard ratios of 2.7 and 1.5, respectively^5^. Despite these epidemiological findings, the immunopathologic interaction between TB and COVID-19 that increases mortality risk remains poorly understood.

The effects of co-infection with *Mycobacterium tuberculosis* (*Mtb*) and SARS-CoV-2 on host immune responses have scarcely been explored in the literature to date. Most published studies have focused on the impact of co-infection on antigen-specific responses to *Mtb* and SARS-CoV-2. These studies have reported that active (clinical) TB limits interferon (IFN)-γ responses to SARS-CoV-2 antigen *in vitro*^6^, that COVID-19 patients have lower levels of *Mtb*-specific CD4+ T cells^7^ and that “latent” TB infection alters humoral responses to SARS-CoV-2 infection^8^ and *vice versa*^9^. A mouse study of *Mtb*/SARS-CoV-2 co-infection demonstrated alteration of the cytokine profile and loss of granulomatous control of *Mtb* in the lung, leading to *Mtb* dissemination^10^. Most recently, immune profiling of TB/COVID-19 co-infected patients revealed significant impairment of antigen-specific responses to the virus and diminished *in vitro Mtb*-specific responses in co-infected patients compared with those with TB-only^11^. Taken together, these data demonstrate an alteration of the ability of the host to respond to and control *Mtb* and/or SARS-CoV-2 in the event of co-infection, prompting further exploration of the underlying immunological pathways.

We previously performed a patient-level meta-analysis of published COVID-19 immune cell signatures on publicly available TB RNA-seq datasets to identify potential immunological hotspots that could exacerbate disease, identifying neutrophil, monocyte, lung macrophage subpopulations, interferon and complement signalling particularly shared between severe COVID-19 and active and subclinical TB^12^. However, a detailed exploration of the responses to genuine co-infection in humans was lacking. We therefore elected to study these responses in human whole blood at single-cell (sc) resolution by subjecting blood samples from four healthy adults to *ex vivo* infection with *Mtb* and/or SARS-CoV-2 for subsequent scRNA-seq at 24 and 96 hours post-infection (p.i.). By defining longitudinal gene expression changes induced by each infectious agent, relative to uninfected samples, and comparing these responses with co-infected samples, we were able to discern unique aspects of the immune response to each pathogen and which responses are exacerbated by co-infection. The present study provides the most comprehensive overview of the immunological interaction between these two globally significant pathogens to date.

## Results

### Pathogen-specific changes in immune cell proportions recovered from peripheral blood emerge over the course of *ex vivo* infection

Peripheral blood collected from four healthy donors, all COVID-19 vaccinated, two with documented PCR-positive mild/moderate COVID-19 and two without were collected on different days and cultured either uninfected or infected with *Mtb*, SARS-CoV-2, or both pathogens simultaneously for 24 and 96 hours. Cells were captured, preserved and stored in HIVE scRNA-seq devices and then batch processed for scRNA-seq library preparation and sequencing. Following acquisition of scRNA-seq data, quality control analysis was first performed on the raw scRNA-seq data acquired from each individual sample from our 8 biological conditions: uninfected, *Mtb*-only, SARS-CoV-2-only (SARS-only) and co-infection, at 2 time points. Each sample was then filtered to exclude cells with <500 genes/transcripts and >15% of reads mapping to mitochondrial genes (indicative of dying cells). This resulted in total cell counts ranging from 367–3,894 (median 2,340), and an overall total of 18,990 cells (Table 1). Normalisation and feature selection were performed prior to data integration, and dimensionality reduction was subsequently performed on the integrated data. *ANPEP*, a gene which encodes alanine aminopeptidase, a receptor known to facilitate the binding and entry of coronaviruses, was the top gene contributing to the first principal component (PC1) of the data (Fig. 1a). In addition to multiple genes involved in macrophage and endothelial cell adhesion and migration, *RAB31*, which encodes a GTPase implicated in the maturation of phagosomes which engulf *Mtb*, was among the top 10 genes contributing to PC1 (Fig. 1a).

**Table 1.** Cell counts for each of the remaining cells belonging to each annotated immune cell population after quality control filtration for each infection condition and timepoint.

| *Cell type* | 24 hours post-infection |  |  |  | 96 hours post-infection |  |  |  |
| --- | --- | --- | --- | --- | --- | --- | --- | --- |
|  | Uninfected | *Mtb*-only | SARS-only | Co-infected | Uninfected | *Mtb*-only | SARS-only | Co-infected |
| *Monocytes* | 2158 | 1106 | 904 | 740 | 2601 | 1464 | 190 | 588 |
| *Neutrophils*<br>(*GM-CSF/ IFN-γ*) | 451 | 158 | 979 | 318 | 317 | 92 | 38 | 60 |
| *Neutrophils (LPS)* | 618 | 297 | 1313 | 820 | 93 | 181 | 17 | 56 |
| *CD8+ T cells* | 304 | 145 | 70 | 87 | 339 | 275 | 47 | 81 |
| *CD4+ T cells* | 247 | 176 | 47 | 138 | 259 | 344 | 46 | 143 |
| *NK cells* | 67 | 63 | 17 | 55 | 55 | 74 | 15 | 42 |
| *HSC*<br>(*G-CSF*) | 34 | 24 | 48 | 28 | 29 | 34 | 9 | 14 |
| *B cells* | 15 | 12 | 2 | 15 | 8 | 15 | 5 | 3 |
| **Total**<br>**(18,990 cells)** | **3894** | **1981** | **3380** | **2201** | **3701** | **2479** | **367** | **957** |

**Fig. 1.**
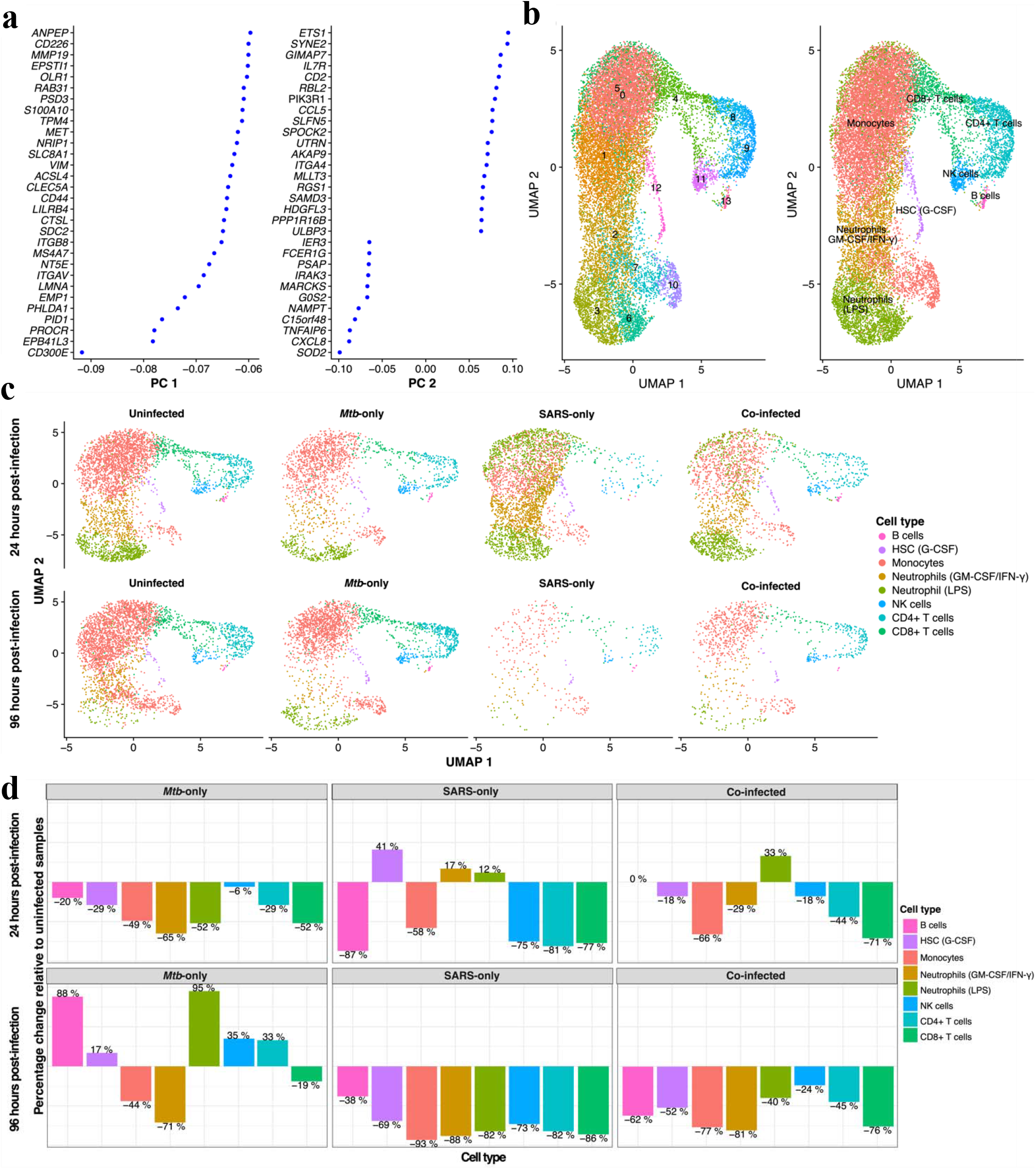
Global changes to the transcriptome of human whole blood in response to single or dual *Mtb*/SARS-CoV-2 infection. **a** The top 30 genes contributing to the first two principal components (PC) identified by performing dimensionality reduction on the integrated scRNA-seq from all eight conditions/timepoints, across four donors. **b** Uniform manifold approximation and projection (UMAP) plot depicting clustering of all 18,990 cells across all eight conditions/timepoints, from four donors, identified by shared nearest neighbour modularity optimisation. Clusters are here grouped according to either the 14 identified clusters (0-13, left) or one of eight distinct immune cell type annotations (right). **c** UMAP plots depicting clustering of cells separated for the four conditions (x-axis) and two timepoints (y-axis) and coloured by the eight distinct immune cell type annotations. Cell number for each condition/timepoint listed in **Table 1**. **d** Bar plots depicting the percentage change in cell count for each of the annotated cell clusters relative to the uninfected samples at the same timepoint, for each infection condition. HSC, haematopoietic stem cell; NK, natural killer; SARS, SARS-CoV-2.

Fourteen clusters were identified based on underlying gene expression differences in the integrated data which were broadly categorised by applying cell type annotation into eight distinct immune cell populations (Fig. 1b). The integrated data were then split by infection condition at each time point to begin comparing cell clusters (Fig. 1c). Yields for each cell type were similar overall between 24 hours and 96 hours p.i. for the uninfected and *Mtb*-only samples, but there was fewer cells recovered in the SARS-only and co-infected samples at 96 h p.i (Fig. 1c, Table 1). SARS-only had the lowest number of cells recovered at this time point, with just 367 remaining after quality filtering (Table 1). Neutrophils made up a greater proportion of the SARS-only and co-infected samples at 24 hours p.i. (68% and 52% of recovered cells, respectively) compared to uninfected and *Mtb*-only (27% and 23%, respectively), with SARS-only having the lowest proportions of CD4+ T cells, CD8+ T cells, B cells and natural killer (NK) cells of all four conditions (Supplementary Fig. 1). This represented a 17% and 12% increase for each of the two identified neutrophil populations in the SARS-only samples, relative to the uninfected samples, while the second population accounted for a 33% increase in the co-infected samples (Fig. 1d). At 96 hours p.i., several immune cell populations were more abundant in the *Mtb*-only samples relative to the uninfected samples (Fig. 1d, Table 1), including B cells (88%), LPS-responding neutrophils (95%), NK cells (35%) and CD4+ T cells (33%). With the exception of B cells, the later 3 cell types were also all higher in the co-infected samples compared with the SARS-only samples at this timepoint, suggesting *Mtb* may be sustaining their viability.

### Distinct immunological pathway enrichment highlights cells responding to infection

For the 14 identified clusters, covering the eight distinct immune cell populations, GO and Reactome biological process (BP) pathway enrichment analyses were performed to determine whether they were associated with distinct immune functions. Pathways were filtered to display the most salient terms in Fig. 2, with the full lists provided in Supplementary Tables 1-2. Clusters 1, 2, 5 and 7 were associated with the greatest number of significantly enriched pathways. While clusters 2 and 5 are both predicted to be neutrophil populations, cluster 2 is characterised by its response to GM-CSF and IFN-γ, whereas cluster 5 is characterised by its greater enrichment of response to LPS. Cluster 5 was found predominantly in the SARS-only and co-infected samples at 24 hours p.i. (Fig. 1c); clusters 3 and 6 were also classified as LPS-responding neutrophils but were not associated with statistically significant enrichment of these pathways at this timepoint. The GM-CSF/IFN-γ-responding neutrophils were associated with the largest number of significantly enriched GO pathways and, as predicted by their annotation, they were defined by response to IFN-γ, production of TNF, cytokine signalling, endocytosis and neutrophil migration (Fig. 2a). Furthermore, this cluster was significantly enriched for Reactome pathways denoting IFN-α/β signalling, Fc-γ receptor-dependent phagocytosis and interleukin signalling (Fig. 2b). The genes implicated in the enrichment of the interleukin signalling term included pro-inflammatory mediators and chemokines *SOD2*, *IRAK1*, *PTAFR*, *IL1B*, *SYK*, *TNFRSF1B*, *CXCL2*, *CSF3R*, *HCK*, *CCL3L1*, *GRB2*, *CCL20*, *CDKN1A*, *CXCL1*, *STAT1*, *CCR1*, *MAP3K8*, *IL1R1*, *CXCL10*, *VEGFA*, *IL1A* and *PTPN12*. Conversely, the LPS-responding neutrophils were exclusively significantly enriched for GO pathways indicating metal ion sequestration and homeostasis (*LCN2*, *FTL*, *TFR2*, *LTF*, *PRNP*, *SLC39A7*, *SLC11A1*, *S100A8* and *S100A9*), regulation of leukocyte cell-cell adhesion, response to chemokine (also confirmed by Reactome pathway analysis, Fig. 2b), and negative regulation of cell adhesion and IFN-γ production (Fig. 2a). Clusters 1, 7 and 10 represent subsets of monocytes, with clusters 1 and 7 both enriched for antigen processing and presentation via MHC class II. Cluster 7 is exclusively enriched for peptide antigen assembly with MHC class II protein complex, whilst Cluster 1 is exclusively enriched for myeloid leukocyte migration, superoxide anion generation and membrane lipid catabolic process and Cluster 10 is exclusively enriched for foam cell differentiation (*ITGAV*, *PPARG*, *MSR1*, *NR1H3*, *IL18*, *ITGB3* and *LPL*).

**Fig. 2.**
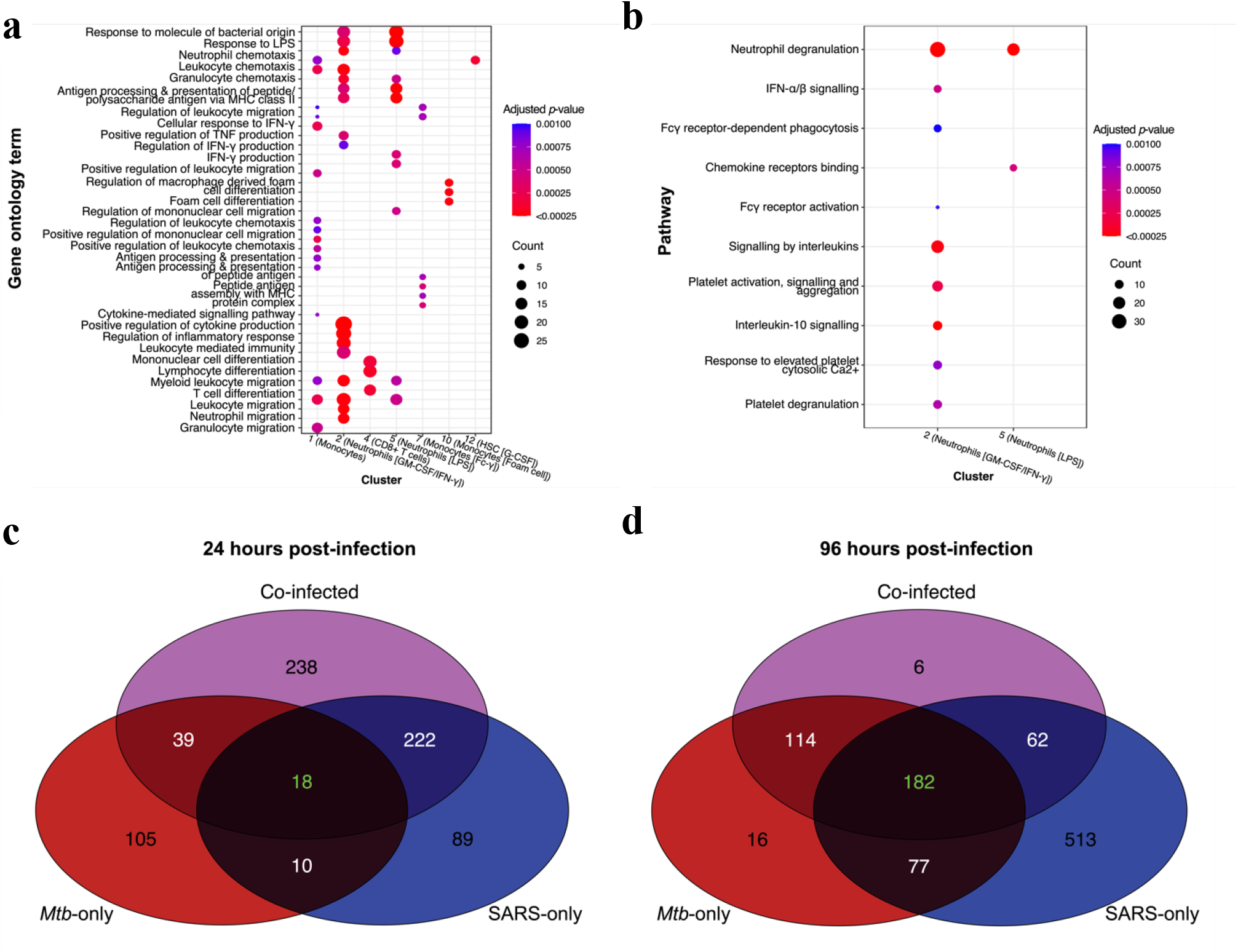
Substantial immune pathway enrichment underpins identified cell clusters and exhibits greatest divergence between conditions at 24 hours post-infection. 14 identified clusters of cells were consolidated to eight immune cell populations based on cluster annotation. Cell marker genes were identified by performing a Wilcoxon Rank Sum tests between each of the eight annotated immune cell populations, irrespective of sample. These differentially expressed gene (DEG) lists were then subjected to a biological process (BP) enrichment analysis using **a** gene ontology (GO) or **b** Reactome pathway annotation. False discovery rate-adjusted 2-sided *p*-values are represented by the shading of data points, while the size corresponds to the number of significant genes (count) implicated in that pathway. A pseudobulk analysis was performed between infection condition at each timepoint by combining cells in each condition and using standard differential expression (DE) methods, with each of the four composite donor IDs as replicates. Significant DEGs were then subjected to a BP GO enrichment analysis and the overlap in GO terms was assessed by venn diagram at **c** 24 hours and **d** 96 hours post-infection. Co-infected, *M. tuberculosis* (*Mtb)* and SARS-CoV-2 (SARS) infected.

### Co-infection demonstrates synergistic global immune pathway enrichment at 24 hours, and greater shared pathways by 96 hours, of infection

To ascertain whether differences in immunological pathway enrichment existed between each infection condition, compared to uninfected, a pseudobulk analysis was performed using donor IDs as biological replicates and performing differential expression analyses across all cell types between conditions. A similar GO and Reactome pathway enrichment analysis (Fig. 2A-B, Supplementary Table 3-4) was then performed on the significant genes identified (Supplementary Table 5-10). Assessing the unique and overlapping significantly enriched terms revealed distinct aspects of the immune response invoked by each infection condition, at both time points. At 24 hour p.i SARS-only and coinfection shared the greatest overlap in terms (222), compared to only 39 commonly enriched terms between *Mtb*-only and co-infection however, this pattern was switched at 96 hours p.i. with *Mtb*-only and co-infection sharing 114 terms compared to only 62 shared terms between SARS-only and co-infection. This demonstrates that the influence of each pathogen on the transcriptome changes during the course of co-infection, with the influence of *Mtb* increasing during co-infection as it replicates over time.

Looking at terms unique to each pathogen, 105 were enriched for *Mtb*-only and 89 for SARS-only at 24 hours p.i.. Of the biologically relevant GO terms for this analysis, certain generic immune cell activation pathways and immune effector processes were uniquely enriched in *Mtb*-only infected cells at 24 hours p.i., while B cell activation (GO:0042113), Fc receptor and STAT signalling (GO:0038094, GO:0097696), and several other cell signalling/response to cytokine GO terms were unique to SARS-only infection (Supplementary Table 3). Amongst the 222 enriched terms shared between SARS-only and co-infection at 24 hours p.i. were αβ T cell activation and differentiation (GO:0046631, GO:0046632), antimicrobial humoral immune response mediated by antimicrobial peptide (GO:0061844), negative regulation of extrinsic apoptotic signalling (GO:2001237), extracellular matrix disassembly (GO:0022617), negative regulation of cell-cell adhesion (GO:0022408), positive regulation of phagocytosis (GO:0050766), regulation of coagulation (GO:0050818), IFN-γ signalling (GO:0034341), IL-1/2/6 production (GO:0032612, GO:0032623, GO:0032635), leukocyte/lymphocyte proliferation (GO:0070661, GO:0046651) and migration (GO:0050900, GO:0072676), antigen processing and presentation (GO:0019882) and defence response to virus (GO:0051607). Amongst, the 39 terms shared between *Mtb*-only and co-infection at 24 hours p.i. included seven related to cell cycle/division, and others relating to endothelial cell migration (GO:0043542) and regulation of cell differentiation (GO:0045595, Supplementary Table 3).

By 96 hour p.i. *Mtb*-only and co-infection were enriched for 114 shared terms including negative regulation of extrinsic apoptotic signalling (GO:2001240) and TNF production (GO:0010804), antigen processing and presentation via MHC class II (GO:0002495), cell death in response to oxidative stress (GO:0036473), iron homeostasis (GO:0006879), endothelial cell migration and proliferation (GO:0043542, GO:0001935), LPS metabolic process (GO:0008653), and reactive nitrogen species (GO:2001057); while SARS-only and co-infection shared 62 terms including myeloid cell differentiation (GO:0030099), pattern recognition receptor/TLR signalling (GO:0002221, GO:0002224) and positive regulation of reactive oxygen species metabolic process (GO:2000379). At 96 hours p.i *Mtb* infection had only 16 unique terms (of its total 389), including WNT signalling (GO:0016055), whereas SARS-only infection had the largest number of enriched terms and the largest number of unique terms at this time (513 of 834 terms, ∼62%, Fig. 2d). However, the unique terms included many variations on terms collectively shared between all three infection conditions at this timepoint (described below) with the additional enrichment of αβ T cell activation and differentiation (GO:0046631, GO:0046632), ephrin receptor signalling pathway (GO:0048013), Fc epsilon receptor signalling (GO:0038095), granulocyte migration (GO:0097530), intrinsic apoptotic signalling (GO:0097193), macrophage activation (GO:0042116), IL-2/6/8 production (GO:0032623, GO:0032635, GO:0032637), monocyte chemotaxis (GO:0002548), and response to IL-1 (GO:0071347) and type I IFN (GO:0034340); many of these terms being shared between SARS-only and co-infection at 24 hours p.i. Thus, over the time course of infection very few of these pathways were unique to SARS-CoV-2 with the majority regulated at one time by both pathogens, just the kinetics varying. Finding the largest enrichment of pathways in the SARS-only infected cells at 96 hours p.i. with this also being the condition with smallest cell recovery, suggests the dominance of the immune response to SARS infection in the absence of *Mtb* co-infection, with the increased cell recovery for co-infected samples perhaps due to survival signals induced by *Mtb* infection.

Next, when comparing how the pathogens interact during co-infection, first comparing the similarity between all three conditions (i.e. terms shared with no synergistic effect of co-infection) there were only 18 collectively shared terms at 24 hour p.i (Fig. 2c), all of which were related to generic biological functions and not specific to infection (Supplementary Table 3). By contrast, co-infection was associated with 238 uniquely enriched terms the largest number across all infection conditions at 24 hours p.i., indicating the synergistic nature of the response to dual infection, identifying terms not induced by either pathogen alone (Fig. 2c); uniquely enriched terms included IFN-γ and TNF production (GO:0034341, GO:0034612), lymphocyte activation/differentiation (GO:0046649, GO:0030098), NK cell-mediated immunity (GO:0002228), T cell signalling (GO:0050852) and positive regulation of viral process (GO:0048525). However by 96 hours p.i. only 6 terms where unique to the co-infection group, indicating limited synergistic pathway enrichment, but there were now 182 terms common between *Mtb*-only, SARS-only and co-infection (Fig. 2d); these included antimicrobial humoral response, B cell activation, T cell differentiation, reactive oxygen species processes, regulation of viral life cycle, lipoprotein metabolic process, tissue remodeling and receptor mediated endocytosis (Supplementary Table 4). Together this shows that *Mtb* and SARS-CoV-2 share a large overlap in immune pathway enrichment which they each induce during infection with early synergism during co-infection.

Finally, at 24 hours p.i there were an additional 10 terms commonly enriched in *Mtb*-only and SARS-only, not shared by co-infection, increasing to 77 commonly enriched terms for the two mono-infections at 96 hours p.i. These terms would be independently but not synergistically regulated by each pathogen, and may include pathways were each pathogen has opposing effects such that during co-infection pathway enrichment is lost.

### Specific differences between infection conditions are most apparent in innate populations and within the first 24 hours of infection

To further examine specific differences and overlapping gene expression between the infection conditions a second pseudobulk analysis was performed, this time at the level of annotated immune cells. For each of the infection conditions, immune cell populations were contrasted against the equivalent cell type in the uninfected group for that timepoint. No significant DEGs were detected in the B cell or haemopoietic stem cell (G-CSF) populations, likely due to the small numbers of cells of this type per donor/condition. The LPS-responding neutrophils at 24 hours p.i. were associated with the greatest number of DEGs across all groups (1,470), with the majority of these induced by co-infection and SARS-only (1,260 total, 434 unique, Supplementary Fig. 2), followed next by monocytes (772 total DEG) and neutrophils (GMCSF) (626 total DEG) at 24 hours p.i. For all cell types with associated differential expression, there were very few DEGs uniquely associated with *Mtb*-only at 24 hours p.i., at which timepoint SARS-only and co-infection induced the greatest DE, while *Mtb*-only and co-infection induced the greatest DE at 96 hours p.i. when SARS-only induced the least DE of all three conditions (Supplementary Fig. 2). This supports a more acute unique transcriptional perturbation associated with SARS-CoV-2 at the initial stages of infection, while *Mtb* induces a response shared by SARS-CoV-2 and co-infection early on which becomes more distinct from SARS-CoV-2 at the later time point after infection is established. The co-infected samples remained highly transcriptionally active throughout both timepoints due to the presence and synergism of both pathogens.

Clustering by DEGs identified for monocyte and neutrophil populations generated the cleanest sample clustering by infection condition and timepoint (Fig. 3a compared with NK, CD4+ and CD8+ T cell populations (Supplementary Fig. 3). At 24hr p.i monocyte expression clustered together for *Mtb*-only and co-infection, whilst co-infection had the most unique monocyte expression at 96 hours p.i., with *Mtb*-only and SARS-only clustering more closely together at this later timepoint. Conversely, patterns of neutrophil gene expression clustering were most distinct at 24 hours p.i at which time GM-CSF neutrophils shared a similar expression pattern between *Mtb*-only and SARS-only infections, whilst LPS neutrophils clustered together for co-infection and SARS-only.

**Fig. 3.**
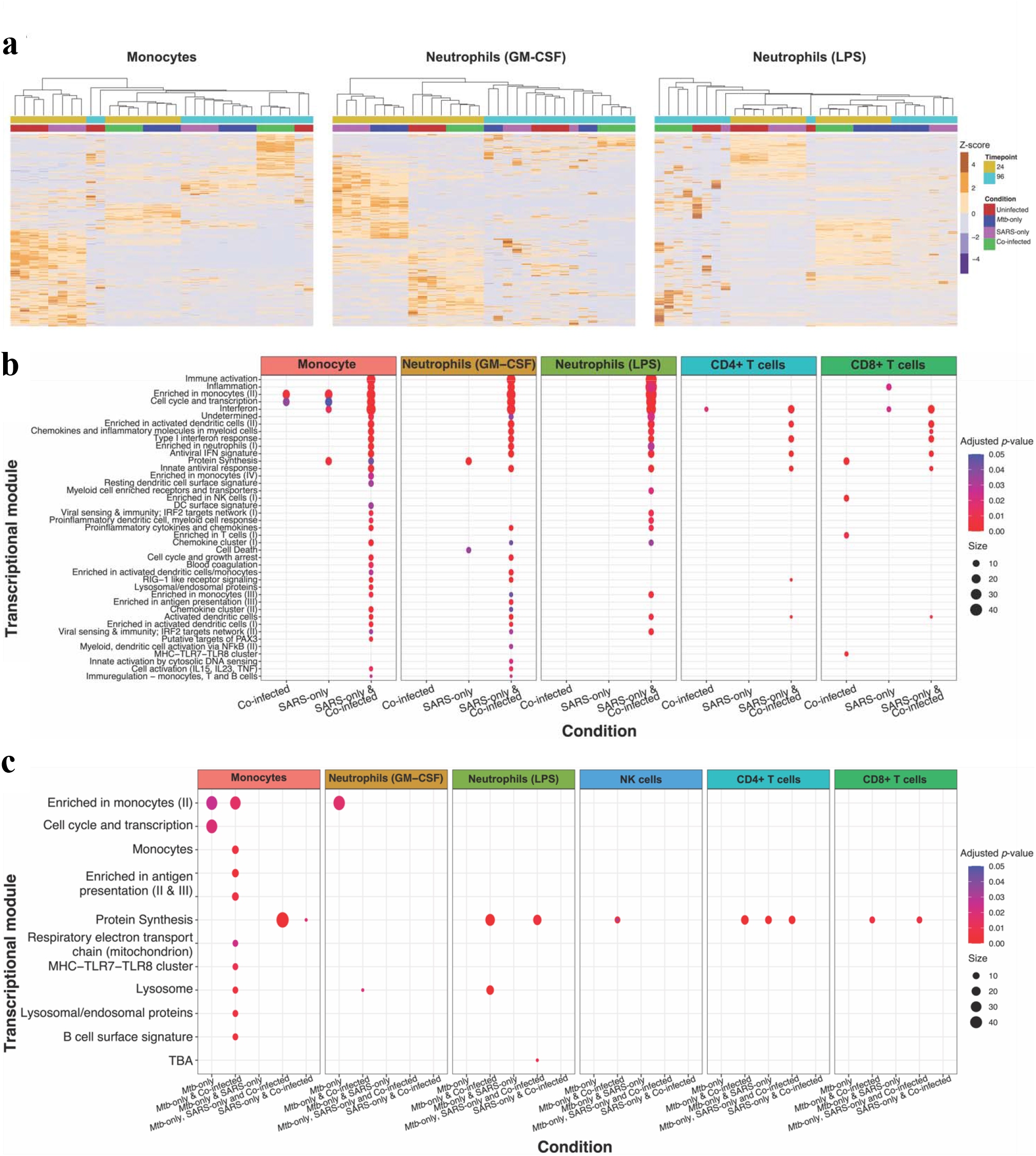
Cell-specific immunological pathway enrichment distinguishes single infection conditions and highlights synergistic activation in co-infected samples. Pseudobulk differential expression (DE) analysis was performed by splitting samples by annotated cell type and comparing these cell types from each infection condition with those of the uninfected samples at each timepoint, using donor IDs as biological replicates. The resultant signatures of significant genes were then used for hierarchical clustering across all samples, represented by the heatmaps in **a**; the heat of the tiles correspond to the row z-score for a particular gene across all samples. Lists of significant genes for each cell type from each condition were subjected to blood transcriptional module analysis to identify significantly enriched pathways associated with immune cells at **b** 24 hours post-infection and **c** 96 hours post-infection. False discovery rate-adjusted 2-sided *p*-values are represented by the shading of data points, while the size corresponds to the number of significant genes implicated in that module.

Finally, the functional enrichment underlying these similarities and differences was assessed at each timepoint. At 24 hours p.i., the majority of enriched pathways were shared by SARS-only and co-infected samples and related to type I IFN responses from monocytes, neutrophils and CD4+ and CD8+ T cells, cell activation (IL-15, IL-23, TNF) of monocytes and neutrophils (GM-CSF, RIG-I like receptor signalling of monocytes, neutrophils (GM-CSF) and CD4+ T cells and viral sensing & immunity; IRF2 targets networks for monocytes and both neutrophil subsets (Fig. 3b, Supplementary Table 11). By comparison, there less cell-specific pathway enrichment at 96 hours p.i., with more enrichment at this timepoint shared between *Mtb-*only and co-infection, or all three infection conditions, with monocytes showing the greatest pathway enrichment, particularly for antigen presentation (II & III) and MHC-TLR7-TLR8 cluster, lysosomes in monocytes and both neutrophils and protein synthesis across all cell types, except neutrophils (GM-CSF) (Fig. 3c, Supplementary Table 12).

## Discussion

In the present study, we expand upon our previous work which identified overlapping transcriptional networks of circulating immune cells isolated from TB and COVID-19 patients ^12^ by delineating host immune cell gene expression programs in response to direct whole blood infection with *Mtb* and SARS-CoV-2 infection. Importantly, we directly compare responses induced by each pathogen alone to what occurs in the context of direct co-infection with both pathogens bringing us closer to an understanding of what is happening at a cellular level during TB/COVID-19 dual presentation. We define the host response pathways underlying differences between immune cell populations reacting to each infection condition across timepoints that appropriately capture innate and adaptive immune cell activity.

Despite the extensive use of bulk RNA-seq to study TB disease mechanisms in the blood of patients, there is but one study that defines host responses to *Mtb* by scRNA-seq in this compartment in which they sequenced PBMCs, excluding granulocytes^13^. There is substantial evidence that neutrophil gene expression is an important marker of TB progression^14–16^, so any comprehensive characterisation of TB responses in human blood must include them. Despite their absence from a large number of blood scRNA-seq datasets, the COVID-19 scRNA-seq studies that did sequence neutrophils as part of whole blood found them to be an important cell type for distinguishing patients on the basis of disease severity^17, 18^. In our study, two neutrophil clusters emerged based on gene expression markers, neutrophils characterised by their response to GM-CSF/IFN-γ and those with enhanced response to LPS. The later were recovered in greater abundance in SARS-only and co-infected blood at 24 hours p.i, whilst only the GM-CSF neutrophils were enriched uniquely in SARS-only infection. Based on GO enrichment analyses, GM-CSF neutrophils appeared to have greater enrichment of IFN-γ and TNF regulation. These are both key cytokines implicated in *Mtb* control^19–21^, so the fact they are not enriched to the same levels during co-infection, although they were more abundant than in *Mtb*-infection alone, suggests *Mtb* is interfering with their regulation by SARS-CoV-2. We previously identified IFN-γ and TNF signalling in the top 20 shared transcriptional pathways in blood of COVID-19 and TB patients and demonstrated *in vitro* that SARS-CoV-2 infected macrophages increase *TNF* and *IFNG* expression when pre-incubated in the inflammatory milieu of *Mtb* infected macrophages, correlating with increased SARS-CoV-2 transcript levels^12^. IFN-γ and TNF have been shown to act synergistically in a mouse model of COVID-19 by increasing inflammatory cell death through a process termed ‘PANoptosis’^22^. Cell death was also exclusively significantly enriched in these neutrophils in the SARS-only samples at 24 hours p.i.. Given the dramatic cell death observed at 96 hours p.i. in SARS-only and co-infected samples, with greater loss of cell recovery in SARS-only compared with co-infected, it is possible that early production of these cytokines from neutrophils may be one of the mechanisms underpinning this. A number of apoptosis regulation GO terms were significantly enriched in SARS-infected samples, with or without *Mtb* co-infection, at 24 hours p.i, including apoptotic cell clearance, extrinsic and intrinsic apoptotic signalling pathways, leukocyte and lymphocyte apoptotic processes and terms associated in the regulation of these pathways (Supplementary Table 3). At 96 hours p.i *Mtb*-only and co-infection shared enrichment of negative regulation of extrinsic apoptosis, which may underpin the increased recovered of cells from co-infected compared to SARS-only infected blood at 96 hours p.i. Caspase 8, the mediator of extrinsic apoptosis, has been identified as a critical factor in triggering inflammatory processes the lead to immunopathology in the lungs of COVID-19 patients^23^. Furthermore, caspase 8-dependent T cell apoptosis has been associated with T cell lymphopenia in severe COVID-19 patients, but this process could be prevented *ex vivo* with the use of pan-caspase inhibitors^24^. Given the critical role of T cells in *Mtb* control, early intervention to prevent T cell death from occurring in co-infected patients may be required to prevent TB disease progression.

Three neutrophil and monocyte cluster signatures were the greatest determinant of differences between the three infection conditions based on hierarchical clustering analyses. This ties in with evidence that innate immune cell gene expression, rather than lymphoid cells, marks the greatest divergence between COVID-19 severity across patients^17^. Monocytes were the cell type associated with the largest number of significantly enriched pathways, the majority of which were common to SARS-only and co-infected samples, specifically those related to immune activation, type I IFN responses and antigen presentation. Monocytes expressing high levels of HLA-DR and type I IFNs were identified in mild COVID-19 patients in the scRNA-seq study exploring myeloid cell dysregulation^17^. It has been shown that the activation status of monocytes are a correlate of COVID-19 prognosis, with inflammatory phenotypes associated with poorer outcomes^25^. Given that certain circulating monocytes have been identified in TB patients which correlate with disease progression, the induction of these activated, proinflammatory monocytes during co-infection may increase the likelihood of TB disease being advanced in co-infected individuals^26^. By 96 hours p.i. the greatest number of significantly enriched pathways was associated with and common to monocytes from *Mtb*-only and co-infected samples and these pathways were mostly related to lysosome and antigen presentation.

The most striking functional response induced by SARS-only/co-infection was the production of IFNs, particularly those classified as type I (α and β). This observation runs counter to some early human studies in which only low levels of type I IFN were measured in COVID-19 patients, irrespective of disease severity^27^, and subsequent studies that suggested little in the way of IFN gene expression in peripheral blood from COVID-19 patients but an early transient wave of IFN-stimulated genes (ISGs)^17, 28^. Mouse studies have indicated that the type I IFN response to SARS-CoV-2 contributes more to immunopathology than viral control^29^. In TB, early expression of ISGs in TB contacts has been linked to disease progression and worsened prognosis^16, 30–32^. Progression from earlier disease states to clinical TB is typically preceded by a wave of ISG expression^16^. However, there is also evidence that type I IFN responses can be protective in TB^33^, and therefore it is likely that the influence of these IFNs on immune cell composition and pathogenesis has a time-dependent effect over the course of TB disease^34^. It is plausible that modulation of the type I IFN response in TB patients with a viral co-infection may alter the balance in such a way as to promote TB disease progression. Over-or under-activation of type I IFN responses is also known to contribute to host pathogenesis for viral infections^35^, so it is equally likely that an existing *Mtb* infection could negatively impact control of SARS-CoV-2 and promote more severe disease manifestations. Our previous patient-level meta-analysis of COVID-19 signatures on TB datasets indicated that IFN gene signatures associated with COVID-19 severity were highly significantly enriched in whole blood of TB progressors and their expression was the strongest correlate of SARS-CoV-2 replication in macrophages^12^, suggesting that the existence of a strong TB IFN signature when SARS-CoV-2 co-infection occurs could be a key determinant of poor outcomes for TB patients.

SARS-only samples had much lower proportions of CD4+ T cell, CD8+ T cell, B cell and NK cell recovery at 24 hours. Longitudinal profiling of lymphocyte subsets in COVID-19 patients showed that this occurs in patients shortly after symptom onset, when severe patients have lower proportions of these cell types than those with mild infections, and they reach a nadir at 4–6 days post symptom onset^36^. It has been reported that TB/COVID-19 co-infected patients have lower absolute lymphocyte counts compared with COVID-19 patients^6^. This did not appear to be the case in our study, although co-infected samples had lower proportions of lymphocytes than *Mtb*-only and uninfected samples at 24 hours. We also observed a strong IFN signature in CD4+ and CD8+ T cells at 24 hours p.i. in the SARS-only and co-infected samples. A scRNA-seq study of peripheral blood responses in severe COVID-19 patients noted that the most commonly upregulated genes in these patients were ISGs based on their DE comparison with healthy controls^37^. At 96 hours p.i protein synthesis was commonly significantly enriched across all three infection conditions in CD4+ and CD8+ T cells. Enrichment of this pathway appeared to be mostly driven by expression of cathepsin genes, including *CTSB* and *CTSD*. These genes encode proteases, mainly found in lysosomes, which have been implicated in *Mtb* survival^38^ and mediating SARS-CoV-2 binding^39^.

The SARS-only samples had a greater proportion of B cells at 96 hours p.i.; several studies have noted an increase in the frequency of plasmablasts in severe COVID-19 patients^28, 40, 41^. However, this cell type was not captured at sufficient levels to perform any meaningful comparisons between groups to ascertain its function. NK cell proportions remained consistent in *Mtb*-only samples, as has been observed in TB patients compared even with TB-HIV co-infected patients^42, 43^. Only samples that were *Mtb*-infected had any significant pathway enrichment in NK cells, with enrichment for protein synthesis, including expression of *CTSD* as well as *PSAP* and *TPT1*, at the 96-hour timepoint. There is evidence that NK cell functionality plays a protective role by combating *Mtb* during HIV infection^44^ that may also apply to other co-infections.

Our study is limited by the fact that we are studying the responses to pathogens in an artificial *ex vivo* infection model, so we do not recapitulate the natural course of infection, including the recruitment of new immune cells to replenish dying ones. Little is known about the mechanisms underlying human immune responses to the initial stages of *Mtb* infection as it is difficult to ascertain when exactly this occurs, while symptoms and diagnostics allow this to be better determined for SARS-CoV-2 infection. Our use of a reagent, TPCK trypsin, to cleave the SARS-CoV-2 spike glycoprotein and allow entry into host cells that do not express or have low levels of ACE2 receptor during initial infection also means that this model is not entirely representative of what occurs *in vivo*. This was done to increase the number of infected cells in our model, over the sort experimental timecourse. Human neutrophils have high levels of ACE2 expression and would be naturally infected and macrophages have been shown to be infected with SARS-CoV-2 in human lung autopsy sections and to be a site for viral replication^45^. We have also shown human macrophages to have increased susceptibility to SARS-CoV-2 infection in the absence of TPCK trypsin addition when incubated in *Mtb* inflammatory milieu^12^. Together this suggests there is the potential for ongoing SARS-CoV-2 infection of immune cells in our model, following TPCK trypsin removal. As not all cells will be infected, this approach allowed us to induce a reliable bystander response in cells that do not take up the virus that could then be compared between conditions. Despite the fact that SARS-CoV-2 transcripts undergo polyadenylation^46^, we were unable to detect any viral transcripts in our count matrices. This may be due to insufficient sequencing depth or the capture efficiency of the scRNA-seq method used. Finally, simultaneous infection with *Mtb* and SARS-CoV-2 is unlikely to occur in most cases. With an established *Mtb* infection prior to SARS-CoV-2 infection more likely to reflect the vast majority of cases, as was observed in the TB/COVID Global Study Group report^4^. Nevertheless, in any co-infected individual there will always be new cells coming into an infection site, so that over time, irrespective of the first infecting pathogen, once both are present then any subsequent sequence of infection can occur.

In summary, we provide the first characterisation of human circulating immune cell gene expression changes resulting from direct *Mtb* and SARS-CoV-2 co-infection. We hope these data will provide a valuable resource for researchers and clinicians to gain insights to better characterise and identify points of potential therapeutic intervention or immunological exacerbation in TB/COVID-19 co-infected patients.

## Materials and Methods

### *Mycobacterium tuberculosis* single cell stock generation

*Mtb* single cell suspension stock for infection were made using *Mtb* lineage 2 clinical strain MRC57, as previously described^12^. Briefly, 10 mL 7H9 (Difco™ Middlebrook 7H9 broth, Becton Dickinson)/ADC (Becton Dickinson) media containing 0.05% Tween-80 (Sigma-Aldrich) were inoculated with *Mtb.* After 10 days growth at 37°C, 1 ml culture was subcultured into 100 mL 7H9/ADC media without Tween-80 and incubated at 37°C for a further 10 days. *Mtb* was pelleted by centrifugation, and then single cell suspensions made by shaking with ∼10 glass beads (2–3 mm, Sigma-Aldrich) with successive washes in PBS and low speed centrifugation. Stocks were made in PBS/5% glycerol and stored at -80°C. One fresh and one frozen aliquot was serially diluted and plated in quadruplicated on three-sector 7H10 (Difco™ Middlebrook 7H10 agar, Becton Dickinson)/ADC plates for colony forming unit (CFU) determination, incubated at 37°C and counted after 10, 14 and 21 days.

### SARS-CoV-2 stock preparation

SARS-CoV-2 stocks for infections were made using isolate VIC001 (24/03/21, obtained from The Peter Doherty Institute for Infection and Immunity, Melbourne, Australia), as previously described^12^. Briefly, vero (CCL-81, ATCC) cells were cultured in Dulbecco’s Modified Eagle Medium (DMEM + 1 g/L D-glucose, L-glutamine and 110 mg/L sodium pyruvate) + 10% heat-inactivated foetal bovine serum (FBS) until confluent. After washing in PBS, 2.5 mL serum-free DMEM containing SARS-CoV-2 MOI of 0.01 (1 x 10^5^ tissue culture infectious dose 50 (TCID50) for ∼1 x 10^7^ cells) and 1 μg/mL TPCK-treated trypsin was added and cells incubated at 37°C with 5% CO_2_ for 30 minutes. 20 mL of serum-free DMEM + TPCK trypsin was added to the flask and incubated at 37°C with 5% CO_2_ for 48 hours or until sufficient cytopathic effect was observed under the microscope. Infection media was centrifuged to pellet debris and 1 ml aliquots of supernatants stored at -80°C. To determine infection stock TCID50, vero cells were seeded in flat-bottomed 96-well plates at1 x 10^4^ cells/well and incubated at 37°C with 5% CO_2_ overnight to achieve confluency. Cells were washed twice with PBS and then cultured in 5-step 1:7 serial dilutions with six replicates/dilution in serum-free DMEM + TPCK and incubated for four days at 37°C with 5% CO_2_. TCID50 values were calculated by scoring wells (positive or negative) for cytopathic effect (CPE) on day four, using the Spearman and Kärber method^47^.

### Blood collection

Acquisition of human blood samples and immunological investigations were approved by the Human Research Ethics Committee at the Walter and Eliza Hall Institute (WEHI HREC #18_09LR and #20/08) and Melbourne Health (RMH69108) as part of the COVID PROFILE study, a longitudinal cohort of convalescent COVID-19 patients and uninfected contacts^48^. Sample and study data were collected and managed using REDCap^49, 50^ electronic data capture tools hosted by Clinical Discovery and Translation, WEHI, Parkville, Victoria, Australia. All subjects provided written informed consent to participate in this study, in accordance with the Declaration of Helsinki. Participants were selected to represent broad demographics of the population: equal age, sex and previous COVID-19 distribution, all COVID-19 vaccinated. Blood was collected in sodium heparin tubes from four healthy participants (2 males [29/47 yrs] and 2 females [23/45 yrs]), three of whom self-identified as being Oceanian (e.g. Australian, Aboriginal, Maori) and one (older female) as People of the Americas (e.g. Hispanic, Brazilian, Mexican, Jamaican). All four had received 2 doses of either Comirnaty (Pfizer/BioNTech) or ChAdOx1-S (Oxford/AstraZeneca). Blood from each participant were divided into eight 500 μL aliquots (approximately 2.5 x 10^6^ cells/aliquot) for two timepoints (24 and 96 hours) and four conditions – uninfected, *Mtb*-infected, SARS-infected and co-infected in 10 ml tubes.

### Blood infections

For SARS-CoV-2 infection, blood samples were pelleted at 300 x *g* for 10 minutes, plasma were transferred to a sterile 2mL tube, cells were resuspended in 1 mL of 2.4 x 10^6^ (TCID50/mL) viral particles of SARS-CoV-2 (MOI 1) in RPMI + TPCK trypsin and incubated at 37°C with 5% CO_2_ for 30 minutes. After incubation, cells were pelleted, infection media removed, and autologous plasma was replaced. For SARS-CoV-2 only infected samples, 500 μL RPMI was then added. *Mtb*-only and co-infected blood samples were infected with 500 μL RPMI containing *Mtb* at 5.5 x 10^4^ CFU/ml. An equivalent volume of RPMI as also added to uninfected samples. All samples were placed on an orbital rotator in a 37°C incubator with 5% CO_2_ for 24 or 96 hours.

### Single-cell RNA-sequencing

Cells were harvested at the appropriate timepoint by pelleting at 300 x *g* for 5 minutes, removing supernatant, resuspending pellets in an appropriate volume of red blood cell lysis solution (0.15 M NH_4_Cl, 0.01 M KHCO_3_, 0.1mM EDTA), incubating for 10 minutes and repeating with an RPMI wash. Cells in a 1 mL RPMI suspension were counted using a Countess automated cell counter (ThermoFisher). Equal numbers of cells from each of the four donors were combined in pairs (one male and one female sample) for each condition and a total of 16 HIVE™ (Honeycomb Biotechnologies) cell capture devices, two for each condition, were loaded with 30,000 total cells according to manufacturer’s instructions for cell preservation and storage at -80°C until ready for further processing. The HIVE™ devices were processed to cDNA, after pooling conditions and timepoints at equimolar concentrations to yield one indexed library for each condition and timepoint (eight in total), according to manufacturer’s instructions (v1 revision A). Final library concentrations were measured using a KAPA Library Quantification Kit (KAPA Biosystems) and profiled on a Tapestation (Agilent) before pooling at equimolar concentrations for Illumina NextSeq 2000 (Illumina) sequencing with custom primers (Honeycomb Biotechnologies), with the base calling and quality scoring performed by the Real Time Analysis (v2.4.6) software.

### Data analysis

The bespoke HIVE™ BeeNet™ pipeline (v1.1, Honeycomb Biotechnologies, https://honeycombbio.zendesk.com/hc/en-us/articles/4408694864283-BeeNet-v1-1-X-Software-Guide) was used to process the raw data into count matrices. First of all, the *make-ref* function was used to generate a human reference genome index from the GRCh38 Homo sapiens reference genome fasta and annotation gtf files (https://ftp.ensembl.org/pub/release-109/fasta/homo_sapiens/). Fastq files were aligned to this index using the BeeNet™ *align* function and transcript count matrices were obtained for each sample. To demultiplex the four donors combined in each sample, cellsnp-lite^51^ was used to pileup expressed alleles in the data and vireo^52^ was subsequently used to assign cells to donors. Donor ID probabilities obtained from these algorithms were used as metadata for downstream analyses requiring biological replicates.

The Seurat pipeline was applied to count matrices for downstream analyses (https://satijalab.org/seurat/). A Seurat object was created for each of the eight samples, combined into a merged Seurat list and subjected to quality control filtration. Features with fewer than 50 genes and 50 unique transcripts were removed and a threshold of <15% mitochondrial reads was used to filter down to the final total cell numbers per sample for analysis. The low gene and transcript thresholds were set to keep cell types with low numbers of expressed genes, particularly neutrophils, in the analysis. Each object in the merged list was subjected to log normalization and variable feature identification, using variance stabilising transformation and 2000 features^53^. Objects were integrated by selecting integration features, finding integration anchors based on these features and using the integration anchors as input for the Seurat *IntegrateData* function^54^. The resultant integrated data object was scaled and subjected to principal component (PC) analysis before graph-based K-nearest neighbour cluster identification was performed. Non-linear dimensionality reduction was performed using uniform manifold approximation and projection (UMAP) on the first 10 dimensions.

Cell type annotation was conducted using SingleR and the Human Primary Cell Atlas Data obtained via the celldex package^55^. Cluster markers were identified by running the Seurat *FindAllMarkers* function using the default Wilcoxon Rank-Sum test to define differentially expressed genes (DEG). Gene ontology (GO) and Reactome pathway enrichment analysis were performed using the identified markers and by invoking the org.Hs.eg.db, AnnotationDbi and clusterProfiler packages^56^. A stringent 2-sided *p*-value cut-off of 0.001 was applied after Benjamini Hochberg adjustment for multiple testing.

To perform pseudobulk differential expression analysis, the Seurat object was first converted to a SingleCellExperiment class. Cluster counts and metadata for each donor were then aggregated and extracted and used to create DESeq2^57^ objects. Counts were transformed for clustering analyses (Supplementary Fig. 4) before running the *DESeq* function. Log_2_ fold-change (LFC) shrinkage and Wald tests were performed for each contrast of interest to obtain results tables for all genes. Those with false-discovery rate (FDR)-adjusted 2-sided *p*-values <0.05 and LFC values >0.58 were deemed significant (Supplementary Table 5-10). GO analyses of pseudobulk data were performed using the clusterProfiler^56^ and tmod^58^ packages.

## Data Availability

The raw fastq files and processed transcript count matrices for this study can be found in the gene expression omnibus database under the accession number GSE232705. The scripts used to analyse these data have been uploaded to GitHub - https://github.com/sheerind-wehi/Mtb_SARS_co-infection

## Data availability

The raw fastq files and processed transcript count matrices for this study can be found in the gene expression omnibus database under the accession number GSEXXXXXX. The scripts used to analyse these data have been uploaded to GitHub - XXXXXX.

## Acknowledgments

The authors thank Honeycomb Biotechnologies Inc. for allowing us to participate in their early access program, Coussens Lab members for feedback, the WEHI PC3 Facility manager Kathryn Davidson for facility support, the WEHI Genomics R&D team, especially Rory Bowden, Daniela Zalcenstein, Daniel Brown and Ling Ling, for their input regarding experimental design and analysis, the Pellegrini Lab at WEHI for providing Vero cells and input into SARS-CoV-2 assays, and Stephen Wilcox of the WEHI Genomics Facility for performing Illumina sequencing of samples. D.S. is supported by WEHI and NHMRC (2020750). A.K.C. is funded by WEHI, philanthropic donors, NHMRC (2020750) and the Australian Respiratory Council. This work was made possible through Victorian State Government Operational Infrastructure Support and Australian Government NHMRC IRIISS. The COVID Profile study was supported by WHO Unity funds and WEHI Philanthropic funds. The funders had no role in study design, data collection and analysis.

## Author Contributions

D.S. and A.K.C. conceived and designed the study. E.E. directed the COVID PROFILE clinical study, arranged blood sample collection, SARS-CoV-2 serotyping, and critically reviewed the manuscript. A.K.C. and E.E funded the study. T.K.P. generated SARS-CoV-2 virus stocks and critically reviewed the manuscript. D.S. generated *Mtb* stocks. D.S. performed blood infection experiments and processed samples for scRNA-seq. D.S. performed the data analysis. D.S. prepared the manuscript with editorial input and revisions from A.K.C. All authors approved the manuscript submission.

## References

1 McQuaid, C. F., Vassall, A., Cohen, T., Fiekert, K. & White, R. G. The impact of COVID-19 on TB: a review of the data. Int J Tuberc Lung Dis 25, 436–446 (2021).

2 Migliori, G. B. et al. Gauging the impact of the COVID-19 pandemic on tuberculosis services: a global study. European Respiratory Journal 58, 2101786 (2021).

3 World Health Organization. Global tuberculosis report 2022. (2022).

4 The TB/COVID-19 Global Study Group. Tuberculosis and COVID-19 co-infection: description of the global cohort. European Respiratory Journal 59, 2102538 (2022).

5 Western Cape Department of Health in collaboration with the National Institute for Communicable Diseases South Africa. Risk Factors for Coronavirus Disease 2019 (COVID-19) Death in a Population Cohort Study from the Western Cape Province, South Africa. Clin Infect Dis 73, e2005–e2015 (2021).

6 Petrone, L. et al. Coinfection of tuberculosis and COVID-19 limits the ability to in vitro respond to SARS-CoV-2. Int J Infect Dis 113 **Suppl 1**, S82–s87 (2021).

7 Riou, C. et al. Relationship of SARS-CoV-2–specific CD4 response to COVID-19 severity and impact of HIV-1 and tuberculosis coinfection. The Journal of Clinical Investigation 131 (2021).

8 Rajamanickam, A. et al. Latent tuberculosis co-infection is associated with heightened levels of humoral, cytokine and acute phase responses in seropositive SARS-CoV-2 infection. Journal of Infection 83, 339–346 (2021).

9 Rajamanickam, A. et al. Effect of SARS-CoV-2 seropositivity on antigen – specific cytokine and chemokine responses in latent tuberculosis. Cytokine 150, 155785 (2022).

10 Hildebrand, R. E. et al. Superinfection with SARS-CoV-2 Has Deleterious Effects on Mycobacterium bovis BCG Immunity and Promotes Dissemination of Mycobacterium tuberculosis. Microbiology Spectrum 10, e03075–03022 (2022).

11 Najafi-Fard, S. et al. Characterization of the immune impairment of tuberculosis and COVID-19 coinfected patients. International Journal of Infectious Diseases (2023).

12 Sheerin, D. et al. Immunopathogenic overlap between COVID-19 and tuberculosis identified from transcriptomic meta-analysis and human macrophage infection. iScience 25, 104464 (2022).

13 Cai, Y. et al. Single-cell transcriptomics of blood reveals a natural killer cell subset depletion in tuberculosis. EBioMedicine 53 (2020).

14 Berry, M. P. R. et al. An interferon-inducible neutrophil-driven blood transcriptional signature in human tuberculosis. Nature 466, 973–977 (2010).

15 Bloom, C. I. et al. Transcriptional blood signatures distinguish pulmonary tuberculosis, pulmonary sarcoidosis, pneumonias and lung cancers. PLoS One 8, e70630 (2013).

16 Scriba, T. J. et al. Sequential inflammatory processes define human progression from M. tuberculosis infection to tuberculosis disease. PLoS Pathog 13, e1006687 (2017).

17 Schulte-Schrepping, J. et al. Severe COVID-19 Is Marked by a Dysregulated Myeloid Cell Compartment. Cell 182, 1419–1440.e1423 (2020).

18 Silvin, A. et al. Elevated Calprotectin and Abnormal Myeloid Cell Subsets Discriminate Severe from Mild COVID-19. Cell 182, 1401–1418.e1418 (2020).

19 Cooper, A. M. et al. Disseminated tuberculosis in interferon gamma gene-disrupted mice. J Exp Med 178, 2243–2247 (1993).

20 Flynn, J. L. et al. An essential role for interferon gamma in resistance to Mycobacterium tuberculosis infection. J Exp Med 178, 2249–2254 (1993).

21 Flynn, J. L. et al. Tumor necrosis factor-alpha is required in the protective immune response against Mycobacterium tuberculosis in mice. Immunity 2, 561–572 (1995).

22 Karki, R. et al. Synergism of TNF-&#x3b1; and IFN-&#x3b3; Triggers Inflammatory Cell Death, Tissue Damage, and Mortality in SARS-CoV-2 Infection and Cytokine Shock Syndromes. Cell 184, 149–168.e117 (2021).

23 Li, S. et al. SARS-CoV-2 triggers inflammatory responses and cell death through caspase-8 activation. Signal Transduction and Targeted Therapy 5, 235 (2020).

24 André, S. et al. T cell apoptosis characterizes severe Covid-19 disease. Cell Death & Differentiation 29, 1486–1499 (2022).

25 Zhang, D. et al. Frontline Science: COVID-19 infection induces readily detectable morphologic and inflammation-related phenotypic changes in peripheral blood monocytes. J Leukoc Biol 109, 13–22 (2021).

26 Lastrucci, C. et al. Tuberculosis is associated with expansion of a motile, permissive and immunomodulatory CD16+ monocyte population via the IL-10/STAT3 axis. Cell Res. 25, 1333–1351 (2015).

27 Hadjadj, J. et al. Impaired type I interferon activity and inflammatory responses in severe COVID-19 patients. Science 369, 718 (2020).

28 Arunachalam, P. S. et al. Systems biological assessment of immunity to mild versus severe COVID-19 infection in humans. Science 369, 1210–1220 (2020).

29 Israelow, B. et al. Mouse model of SARS-CoV-2 reveals inflammatory role of type I interferon signaling. J Exp Med 217 (2020).

30 Zak, D. E. et al. A blood RNA signature for tuberculosis disease risk: a prospective cohort study. Lancet 387, 2312–2322 (2016).

31 Singhania, A. et al. A modular transcriptional signature identifies phenotypic heterogeneity of human tuberculosis infection. Nat Commun 9, 2308 (2018).

32 Esmail, H. et al. Complement pathway gene activation and rising circulating immune complexes characterize early disease in HIV-associated tuberculosis. Proceedings of the National Academy of Sciences 115, E964–E973 (2018).

33 Zhang, G. et al. A proline deletion in IFNAR1 impairs IFN-signaling and underlies increased resistance to tuberculosis in humans. Nature Communications 9, 85 (2018).

34 Desvignes, L., Wolf, A. J. & Ernst, J. D. Dynamic roles of type I and type II IFNs in early infection with Mycobacterium tuberculosis. J Immunol 188, 6205–6215 (2012).

35 Mesev, E. V., LeDesma, R. A. & Ploss, A. Decoding type I and III interferon signalling during viral infection. Nat Microbiol 4, 914–924 (2019).

36 Liu, J. et al. Longitudinal characteristics of lymphocyte responses and cytokine profiles in the peripheral blood of SARS-CoV-2 infected patients. eBioMedicine 55 (2020).

37 Wilk, A. J. et al. A single-cell atlas of the peripheral immune response in patients with severe COVID-19. Nat. Med. 26, 1070–1076 (2020).

38 Pires, D. et al. Role of Cathepsins in Mycobacterium tuberculosis Survival in Human Macrophages. Sci Rep 6, 32247 (2016).

39 Zhang, Q. et al. Molecular mechanism of interaction between SARS-CoV-2 and host cells and interventional therapy. Signal Transduction and Targeted Therapy 6, 233 (2021).

40 Mathew, D. et al. Deep immune profiling of COVID-19 patients reveals distinct immunotypes with therapeutic implications. Science 369 (2020).

41 Kuri-Cervantes, L., et al. Comprehensive mapping of immune perturbations associated with severe COVID-19. Sci Immunol 5 (2020).

42 Ardain, A. et al. Group 3 innate lymphoid cells mediate early protective immunity against tuberculosis. Nature 570, 528–532 (2019).

43 Roy Chowdhury, R., et al. A multi-cohort study of the immune factors associated with M. tuberculosis infection outcomes. Nature 560, 644–648 (2018).

44 Allen, M. et al. Mechanisms of Control of Mycobacterium tuberculosis by NK Cells: Role of Glutathione. Front Immunol 6, 508 (2015).

45 Sefik, E. et al. Inflammasome activation in infected macrophages drives COVID-19 pathology. Nature 606, 585–593 (2022).

46 Chang, J. J., et al. Long-Read RNA Sequencing Identifies Polyadenylation Elongation and Differential Transcript Usage of Host Transcripts During SARS-CoV-2 In Vitro Infection. Front Immunol 13, 832223 (2022).

47 Ramakrishnan, M. A. Determination of 50% endpoint titer using a simple formula. World J Virol 5, 85–86 (2016).

48 Emily, M. E., et al. Cohort Profile: A longitudinal Victorian COVID-19 cohort (COVID PROFILE). medRxiv, 2023.2004.2027.23289157 (2023).

49 Harris, P. A. et al. Research electronic data capture (REDCap)--a metadata-driven methodology and workflow process for providing translational research informatics support. J Biomed Inform 42, 377–381 (2009).

50 Harris, P. A. et al. The REDCap consortium: Building an international community of software platform partners. J Biomed Inform 95, 103208 (2019).

51 Huang, X. & Huang, Y. Cellsnp-lite: an efficient tool for genotyping single cells. Bioinformatics 37, 4569–4571 (2021).

52 Huang, Y., McCarthy, D. J. & Stegle, O. Vireo: Bayesian demultiplexing of pooled single-cell RNA-seq data without genotype reference. Genome Biology 20, 273 (2019).

53 Hafemeister, C. & Satija, R. Normalization and variance stabilization of single-cell RNA-seq data using regularized negative binomial regression. Genome Biol 20, 296 (2019).

54 Stuart, T. et al. Comprehensive Integration of Single-Cell Data. Cell 177, 1888–1902.e1821 (2019).

55 Aran, D. et al. Reference-based analysis of lung single-cell sequencing reveals a transitional profibrotic macrophage. Nature Immunology 20, 163–172 (2019).

56 Wu, T. et al. clusterProfiler 4.0: A universal enrichment tool for interpreting omics data. The Innovation 2 (2021).

57 Love, M. I., Huber, W. & Anders, S. Moderated estimation of fold change and dispersion for RNA-seq data with DESeq2. Genome Biology 15, 550 (2014).

58 Zyla, J. et al. Gene set enrichment for reproducible science: comparison of CERNO and eight other algorithms. Bioinformatics 35, 5146–5154 (2019).

